# Associations between blood cholinesterase activity and postoperative basal forebrain cholinergic system atrophy: Results from the BioCog cohort study

**DOI:** 10.1101/2025.09.05.25335162

**Authors:** Florian Lammers-Lietz, Maria Heinrich, Kai Kappert, Georg Winterer, Claudia Spies, Anika Müller

## Abstract

**OBJECTIVE:** Structural and functional metabolism are closely linked in the cholinergic system, and associations between cholinergic neurotransmission and atrophy of cholinergic nuclei, i.e., the nucleus basalis of Meynert (NBM), have been described for neurodegenerative conditions. Perioperative drug administration significantly impacts cholinergic neurotransmission, as shown by alterations in cholinesterase (ChE) activity of patients undergoing anaesthesia and may contribute to perioperative disorders of cognition. In this work, we hypothesised that there is an association between perioperative ChE activity and postoperative NBM atrophy and studied its potential role in postoperative delirium (POD) and cognitive dysfunction (POCD).

**METHODS:** We studied a subsample from the BioCog cohort study. N=170 cognitively healthy (MMSE≥24) patients ≥65 years provided blood samples for ChE activity measurement before elective surgery and on the first postoperative day, as well as magnetic resonance imaging data before surgery and at follow-up three months after surgery for volumetry of the NBM. Patients were screened for delirium until the 7^th^ postoperative day. POCD was assessed three months after surgery and was defined according to the reliable change index. The data were analysed in linear regression models of postoperative NBM volume adjusted for age, sex, MMSE score and preoperative NBM volume.

**RESULTS:** Lower postoperative activity of butyryl-ChE was associated with lower baseline-adjusted NBM volume, which was driven by the perioperative change in butyryl-ChE activity rather than preoperative activity. Significant associations were not observed for preoperative butyryl-ChE activity or pre- or postoperative acetyl-ChE activity. Postoperative NBM volumes were not altered in patients with POD or POCD, and associations between butyryl-ChE and postoperative NBM volume were not altered in patients with POD or POCD compared with patients without postoperative cognitive disorder.

**DISCUSSION:** We observed an association between plasma butyryl-ChE activity and postoperative NBM atrophy. Our findings may point toward a role of surgery and anaesthesia for neurodegenerative diseases and toxic encephalopathies. However, the involvement of this mechanism in postoperative cognitive disorders could not be proven.

## 1 Introduction

Acetylcholinesterase (AChE) is essential for neuronal function because it terminates the action of the neurotransmitter acetylcholine (ACh). It may be involved in neurito- and synaptogenesis, cell adhesion, dopaminergic neurotransmission, haematopoiesis, thrombopoiesis, and even amyloid fibre assembly (Soreq & Seidman, 2001). In addition to its expression in neuronal tissue, AChE is expressed in muscles, lung, retina, myometrium, endothelial and glial cells, blood cells and erythrocytes (Richbart, Merritt, Nolan, & Dasgupta, 2021). While AChE has strong specificity for ACh as a substrate, butyrylcholinesterase (BuChE, also referred to as pseudocholinesterase) hydrolyses a variety of both choline and non-choline esters (Darvesh, Hopkins, & Geula, 2003). Except for the brain, BuChE activity exceeds AChE activity in most tissues, e.g., the intestines, the liver, the blood, and the heart (Li et al., 2000). In the brain, BuChE is expressed predominantly by glial cells, although neurons in the hippocampus, amygdala and thalamus also synthesise BuChE (Darvesh et al., 2003; Johnson & Moore, 2012). The exact function of BuChE is poorly understood, and potential roles of BuChE as a back-up or coregulatory enzyme in cholinergic neurotransmission, or as a scavenger molecule for natural AChE-inhibitors have been suggested. Furthermore, BuChE is thought to be involved in nervous system development, and plaque formation in Alzheimer’s disease (Darvesh et al., 2003).

The nucleus basalis of Meynert (NBM) is the main source of cortical ACh (Mesulam & Geula, 1988), and plays a central role in cognitive function, especially in neurodegenerative diseases, such Alzheimer’s disease (AD) (M. Grothe, Heinsen, & Teipel, 2012; M. Grothe et al., 2010; M. J. Grothe et al., 2014; Pereira et al., 2020; Rogozinski et al., 2022). It is unknown whether dysregulation of AChE or BuChE activity directly affects the NBM in neurodegenerative disease, but in the past, several pathways have been described by which dysregulation of cholinesterase (ChE) activity may contribute to atrophy of the NBM: ChE activity may regulate systemic inflammation (Hofer et al., 2008; Ofek et al., 2007; Pollak et al., 2005; Shenhar-Tsarfaty, Berliner, Bornstein, & Soreq, 2014), which was found to be associated with NBM atrophy (M. Heinrich et al., 2024b). In addition, ChE activity may influence neurotrophic factors that impact NBM atrophy. Cholinergic neurons of the basal forebrain seem to be particularly responsive to nerve growth factor (NGF) signalling (Cellerino, 1996; Holtzman et al., 1995). Conversely, ACh is an inducer of NGF release; hence, stable cholinergic neurotransmission may contribute to cholinergic neuron survival (French, Humby, Horner, Sofroniew, & Rattray, 1999; Mufson, Bothwell, Hersh, & Kordower, 1989). In line with this theory, reduced atrophy of the basal forebrain cholinergic system has previously been described in the interventional group of a randomised clinical trial on the effectiveness of the AChE-inhibitor donepezil (Cavedo et al., 2017).

On the other hand, blood ChE activity is utilised as a biomarker for ChE-inhibitor poisoning, e.g. with chemical warfare agents (Thiermann et al., 2007), insecticides (Kapeleka, Sauli, Sadik, & Ndakidemi, 2019; Peiris-John, Ruberu, Wickremasinghe, Smit, & van der Hoek, 2002; Vikkey et al., 2017), lead (Ademuyiwa et al., 2007), laboratory chemicals (Li et al., 2000), and drugs (Alygizakis et al., 2016; Proctor, Petrie, Barden, Arnot, & Kasprzyk-Hordern, 2019). ChE inhibition and possibly overshooting cholinergic neurotransmission have been found to be associated with incident neurodegenerative disease and brain atrophy (Chao, Abadjian, Hlavin, Meyerhoff, & Weiner, 2011; Chao, Rothlind, Cardenas, Meyerhoff, & Weiner, 2010; Chuang, Su, Lin, & Kao, 2017; Wang, Cockburn, Ly, Bronstein, & Ritz, 2014; Yamasue et al., 2007; Yan, Zhang, Liu, Shi, & Yan, 2018). The molecular mechanisms by which cholinergic neurotransmission triggers neuronal death are unknown, but may include immunosuppression with impaired defence mechanisms against tissue damage (Ofek et al., 2007; Pollak et al., 2005), the impact of ACh depletion on neuronal energy metabolism (Siegel, 2006) or actions unrelated to the hydrolysis of ACh (Soreq & Seidman, 2001).

POD is a common complication especially in older patients undergoing anaesthesia for surgery (Aldecoa et al., 2024; Aldecoa et al., 2017). In addition to acute cognitive deterioration, POD is associated with long term cognitive impairment (Kunicki et al., 2023; Teipel et al., 2018). Previous studies have shown that dysregulation of peripheral cholinesterase activity in patients undergoing anaesthesia and surgery is associated with postoperative delirium (POD) (Bosancic et al., 2022; Muller et al., 2019) and, vice versa, that the regulation of ChE activity may mediate the anti-delirogenic effects of dexmedetomidine (Jacob et al., 2023; van Norden et al., 2021). On the other hand, studies on involvement of the central cholinergic nervous system have been inconclusive: Preoperative AD-like cortical atrophy (Racine et al., 2017), but not preoperative NBM atrophy in particular, has been observed in patients with POD(Florian Lammers-Lietz et al., 2024). In contrast, preoperative tissue alterations in the basal forebrain cholinergic system were associated with POD in two studies (Cavallari et al., 2016; F. Lammers-Lietz et al., 2024). It is also debatable whether cholinergic neurons are particularly vulnerable to dysregulated cholinergic neurotransmission in POD (Cavallari et al., 2017), although AD-like cortical atrophy and mineralization of cholinergic nuclei in the basal forebrain have been found to be associated with postoperative cognitive decline (F. Lammers-Lietz et al., 2024; Racine et al., 2020).

To better understand the interactions among peripheral cholinergic function, the structural integrity of the central cholinergic system and their relevance for POD and postoperative cognitive dysfunction (POCD), we studied the associations between perioperative plasma ChE activity and postoperative NBM atrophy in a cohort of cognitively healthy patients ≥65 years of age undergoing elective surgery.

We hypothesise that perioperative plasma ChE activity is associated the with baseline-adjusted postoperative NBM volume. We further analysed associations of POD and POCD incidence with postoperative NBM atrophy and their modulatory effects on the association between plasma ChE activity and postoperative NBM volume change.

## 2 Methods

### 2.1 Study design

BioCog (www.biocog.eu, clinicaltrials.gov: NCT02265263 on Oct 14 2014) is a prospective observational cohort study with the aim of identifying POD and POCD risk factors. The methodology of the BioCog study has been published previously (Bosancic et al., 2022; M. Heinrich et al., 2024a; Florian Lammers-Lietz et al., 2024; Lammers-Lietz et al., 2022; F. Lammers-Lietz et al., 2024; Winterer, Androsova, Bender, Boraschi, Borchers, Dschietzig, Feinkohl, Fletcher, Gallinat, Hadzidiakos, Haynes, Heppner, Hetzer, Hendrikse, Ittermann, Kant, Kraft, Krannich, Krause, Kuhn, Lachmann, van Montfort, Muller, Nurnberg, Ofosu, Pietsch, Pischon, Preller, Renzulli, Scheurer, Schneider, Slooter, Spies, Stamatakis, Volk, Weber, Wolf, Yurek, Zacharias, & BioCog, 2018).

### 2.2 Participants

Male and female patients were enrolled in two tertiary care centres at the Charité– Universitätsmedizin Berlin, Germany, and the University Medical Center Utrecht, Netherlands. Patients aged ≥65 years presenting for elective surgery with an expected duration >60min were included. Patients meeting one of the following criteria were excluded:

− positive screening for preexisting major neurocognitive disorder defined as a Mini-Mental Status Examination (MMSE) score ≤23 points
− any condition interfering with neurocognitive assessment
− unavailability for follow-up assessment
− accommodation in an institution due to official or judicial order
− inability to provide informed consent

ChE activity measurements were only conducted at the Berlin study site.

### 2.3 Study procedures

Consenting patients underwent a preoperative baseline assessment including MRI, blood sampling, cognitive testing, and collection of clinical data. Blood sampling was repeated on the first postoperative day. Patients were screened for postoperative delirium until the seventh postoperative day. After three months, patients were invited to a follow-up assessment including MRI and cognitive testing.

### 2.4 Cholinesterase activity

BuChE and Hb-adjusted AChE-activity were measured before surgery and on the first postoperative day. As described previously (Bosancic et al., 2022; Maria Heinrich et al., 2020; Jacob et al., 2023; Muller et al., 2019), ChE activity was measured in 10µL of whole blood with a validated photometric point-of-care testing device in accordance with the manufacturer’s instructions (ChE check mobile, Securetec Detektions-Systeme AG, Neubiberg, Germany).

### 2.5 MRT

For T1-weighted structural image acquisition, an MPRAGE sequence (magnetization prepared rapid gradient echo in 192 sagittal slices, FOV: 256·256mm², voxel size: 1·1mm² at 1mm slice thickness, TR: 2500ms, TE: 4.77ms, 7° flip angle) was run on a 3T Magnetom Trio MR scanner (Siemens) equipped with a 32-channel head coil at the Berlin Center for Advanced Neuroimaging (BCAN). The complete neuroimaging protocol is described elsewhere (Winterer, Androsova, Bender, Boraschi, Borchers, Dschietzig, Feinkohl, Fletcher, Gallinat, Hadzidiakos, Haynes, Heppner, Hetzer, Hendrikse, Ittermann, Kant, Kraft, Krannich, Krause, Kuhn, Lachmann, van Montfort, Muller, Nurnberg, Ofosu, Pietsch, Pischon, Preller, Renzulli, Scheurer, Schneider, Slooter, Spies, Stamatakis, Volk, Weber, Wolf, Yurek, Zacharias, & Consortium, 2018).

#### 2.5.1 Image processing

SPM12 (http://www.fil.ion.ucl.ac.uk/spm/software/spm12/) in a MATLAB environment (The Mathworks. Inc. Natick. MA) was used for volumetric MRI analysis with additional use of the log_roi_batch extension by Adrian Imfeld (http://www.aimfeld.ch/neurotools/neurotools.html). MR images were partitioned into gray and white matter as well as cerebrospinal fluid using the SPM12 segmentation routine. For segmentation of the basal forebrain cholinergic system (BFCS), including the NBM, a previously described probabilistic map was used (Zaborszky et al., 2008). At each voxel in this map, the probability of presence of cholinergic cells was ≥40% on the basis of ten post-mortem brain specimen from which this map was derived. Both the tissue map and the segmented patient data were mapped onto a common template via the DARTEL (Diffeomorphic Anatomical Registration using Exponentiated Lie algebra) flow fields implemented in SPM12. These deformations were applied to the probabilistic map of the basal forebrain resulting in individual labelling of cholinergic subregions in a participant’s brain scan.

The BFCS tissue map differentiates between four subregions (Ch1/2, Ch3, Ch4 and Ch4p) according to a modified version of Mesulam’s nomenclature of the cholinergic system. NBM volume was calculated as the sum of voxels in the subregions Ch4 and Ch4p.

### 2.6 Assessment of POD

Independent of routine hospital procedures, POD screening was started in the recovery room and repeated twice per day at 8:00 am and 7:00 pm (±1h) up to seven days after surgery by or under supervision of a study physician. POD was defined according to the DSM-5 criteria and assessed by prospective screening with three validated tools that were recorded at each visit in accordance with current guidelines (Aldecoa et al., 2024; Aldecoa et al., 2017), and chart review to mitigate the known tendency of physicians to underdiagnose POD. Patients were considered delirious if at least one of the following criteria was met:

- ≥2 points on the Nursing Delirium Screening Scale (Nu-DESC),
- positive Confusion Assessment Method (CAM) score on a general ward,
- positive CAM for the Intensive Care Unit (CAM-ICU) score in an intensive care unit,
- chart review showing descriptions of delirium.

### 2.7 Assessment of POCD

Neuropsychological testing took place at baseline before surgery, and three months after surgery. To adjust for natural variability in cognitive performance and learning effects in repeated cognitive testing, POCD was defined according to the Reliable Change Index (RCI) with the ISPOCD criteria proposed by Rasmussen (Rasmussen et al., 2001). The RCI is the patient’s change in performance at a cognitive test parameter after surgery compared with baseline performance in relation to the mean change in a nonsurgical control group. POCD was diagnosed in patients with an RCI score below -1.96 in at least two single cognitive test parameters (i.e., showing a more severe performance decline than approximately 97.5% of the reference population) and/or a compound RCI averaged over all single RCIs in the follow-up assessment three months after surgery. 114 nonsurgical control participants without indication for surgery but otherwise identical in- and exclusion criteria were recruited from outpatient clinics, primary care facilities, nursing homes and via calls during public talks(Feinkohl et al., 2020) and performed consecutive neuropsychological testing at baseline and three months to adjust for natural learning effects in cognitive testing (Rasmussen et al., 2001).

#### 2.7.1 Neuropsychological testing

Testing was performed by trained study assistants in accordance with a standard operating procedure, which was approved by two neuropsychologists.

The following cognitive test parameters have been used for the calculation of POCD: mean correct latency from the Simple Reaction Time (SRT, processing speed), the number of correctly remembered items in the free recall task (VRM free recall) as well as the number of correctly recognised items after delay on the Verbal Recognition Memory test (VRM recognition, verbal memory), span length in the Spatial Span task (SSP, working memory), the first trial memory score from the Paired Associate Learning test (PAL, visual memory) as implemented in the CANTAB test battery (RRID:SCR_003001), and the completion times of the Trail-Making-Test-B (TMT-B, executive functions) and the Grooved Pegboard test (GPT, fine motor skills). We selected these tests because of their moderate-to-good retest-reliability in the control group (intraclass coefficient between baseline and three months ≥0.75 on the basis of a mean of multiple measurements, absolute-agreement, 2-way mixed-effects models) for this purpose(Feinkohl et al., 2020). Details on single cognitive tests are given elsewhere(Lammers et al., 2018).

Two independent assessors checked the data on plausibility including the review of free-text entries of research team members. When data for a participant were incomplete, missing values were imputed. If the data were missing due to impairment of concentration or poor understanding of test instructions according to the administering researcher, the missing data were replaced with the worst performance value of the entire patient group, under the assumption that missingness reflects inability to perform the test due to cognitive decline. When values were missing at random, e.g., due to technical difficulties or environmental disturbances, random forest imputation was applied to replace missing values. Data were not imputed when neuropsychological testing was missing completely. The missForest package for R was used for imputations (Stekhoven & Bühlmann, 2012).

#### 2.7.2 Calculation of the reliable change index

Prior to calculation, SRT, GPT and TMT-B were log-transformed and sign-reversed to achieve an approximate normal distribution and a correspondence of higher scores with better cognitive performance. For each cognitive test parameter, the corresponding RCI was calculated as RCI=(ΔX-ΔX_c_)/sd_ΔXc_ as implemented in the POCDr package for R Statistical Software (https://github.com/Wiebachj/POCDr) (Spies et al., 2021). ΔX refers to the difference in test scores after surgery compared with baseline and ΔX_c_ refers to the mean test score difference between the corresponding measurement time points in the non-surgical control group. RCI was normalised to the standard deviation (sd) of the mean differences in the control group (sd_ΔXc_). The compound RCI for each patient was defined as the sum of all RCIs in relation to the standard deviation of the sum of RCIs in the control group (RCI_c_): compound RCI=Σ(RCI)/sd_Σ(RCIc)_.

### 2.8 Statistical analysis

All the statistical analyses were conducted in R (RRID:SCR_001905). In all the models, the postoperative NBM volume at follow-up after three months was treated as the dependent variable. To model postoperative atrophy rather than postoperative volume, preoperative NBM volume was included as an independent variable (referring to baseline-adjusted postoperative volume as the dependent variable), i.e., a lower baseline-adjusted postoperative NBM volume indicates increased NBM atrophy. Age, sex and MMSE score at study enrolment were included as covariates of no interest.

#### 2.8.1 Associations between ChE activity and NBM atrophy

To study the associations between preoperative ChE activity and NBM atrophy, preoperative AChE and BuChE activity were included as independent variables of interest.

To study the associations between postoperative ChE activity and NBM atrophy, BuChE and AChE activity on the first postoperative day were the independent variables of interest. To further disentangle the effects of preoperative BuChE activity and postoperative BuChE activity changes in response to anaesthesia and surgery, we included preoperative BuChE activity and postoperative BuChE change measured as the difference between postoperative and preoperative activity in the model.

The level of significance was set to p≤0.05 and adjusted for four independent tests (two independent ChE activities measured at two independent time points) with Bonferroni’s method, yielding p≤0.0125.

In the supplementary material, we also provide results for anaemia-adjusted AChE activity and provide albumin-adjusted models to account for liver function as a confounder of BuChE activity. To determine if our findings were restricted to atrophy of the cholinergic system, we repeated the analysis with replacement of NBM volume by global brain volume to assess confound by global brain atrophy.

#### 2.8.2 Associations between ChE activity and NBM atrophy in patients with POD and POCD

To study effects of POD and POCD on postoperative NBM atrophy, these variables were included as independent variables of interest. The overall level of significance was adjusted for two independent tests of associations with POD or POCD, yielding p≤0.025.

To study the modulatory effect of POD and POCD on the associations between ChE activity and NBM atrophy, we calculated the interactions between POD or POCD and ChE activity. On the basis of the observed associations between ChE activity and NBM atrophy, we restricted these analyses to interactions between postoperative BuChE and postoperative change in BuChE activity. AChE and preoperative BuChE activity were not considered in these models. In the analysis of postoperative BuChE activity change in POD, we observed one potentially influential outlier in partial regression plots and report results after removal of this patient from the dataset. The overall level of significance was adjusted for two independent tests of interactions with POD or POCD, yielding p≤0.025.

#### 2.8.3 Sample size estimation

The sample size estimation for the primary aim of the BioCog study is outlined in detail elsewhere(Florian Lammers-Lietz et al., 2024; Winterer, Androsova, Bender, Boraschi, Borchers, Dschietzig, Feinkohl, Fletcher, Gallinat, Hadzidiakos, Haynes, Heppner, Hetzer, Hendrikse, Ittermann, Kant, Kraft, Krannich, Krause, Kuhn, Lachmann, van Montfort, Muller, Nurnberg, Ofosu, Pietsch, Pischon, Preller, Renzulli, Scheurer, Schneider, Slooter, Spies, Stamatakis, Volk, Weber, Wolf, Yurek, Zacharias, & BioCog, 2018). In total, a sample size of N=1200 was planned. Here, we present a secondary analysis of the data, for which sample size calculation has been performed. The final sample size presented here was arrived by inclusion of all patients who provided ChE measurement and NBM volumetry.

## 3 Results

### 3.1 Sample

A total of 5294 patients were screened in Berlin, of whom 747 patients were included. 61 patients dropped out, yielding 686 patients who underwent baseline assessments. Of these, 516 were excluded because of either missing longitudinal NBM or ChE measurements. In the final sample of 170 patients, N=13 and N=15 patients did not provide AChE and BuChE levels on the first postoperative day. Figure 1 displays the STROBE chart. Table 1 describes the cohort of N=170 patients.

**Figure 1:**
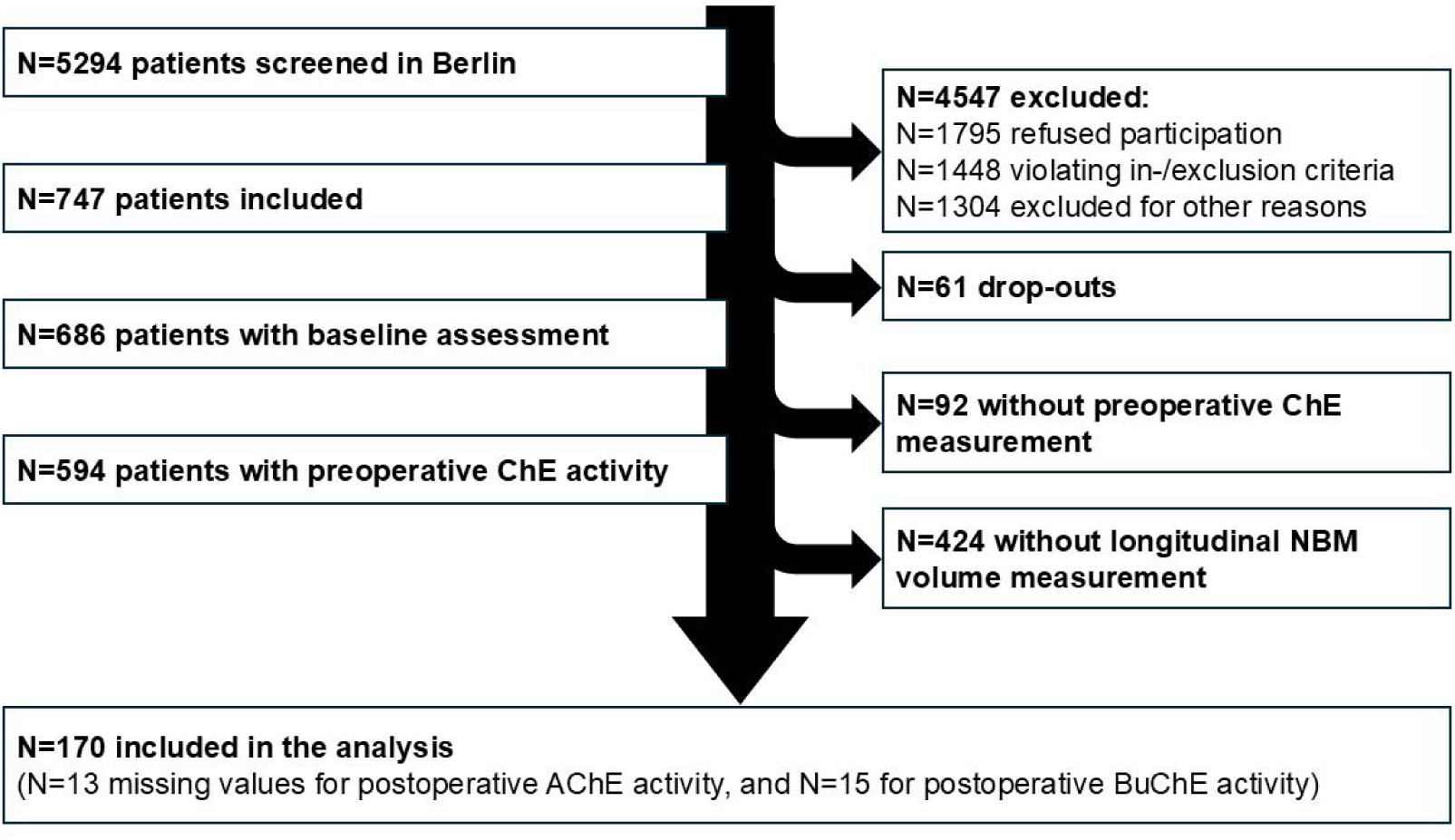
Patient flow chart.

**Table 1:**
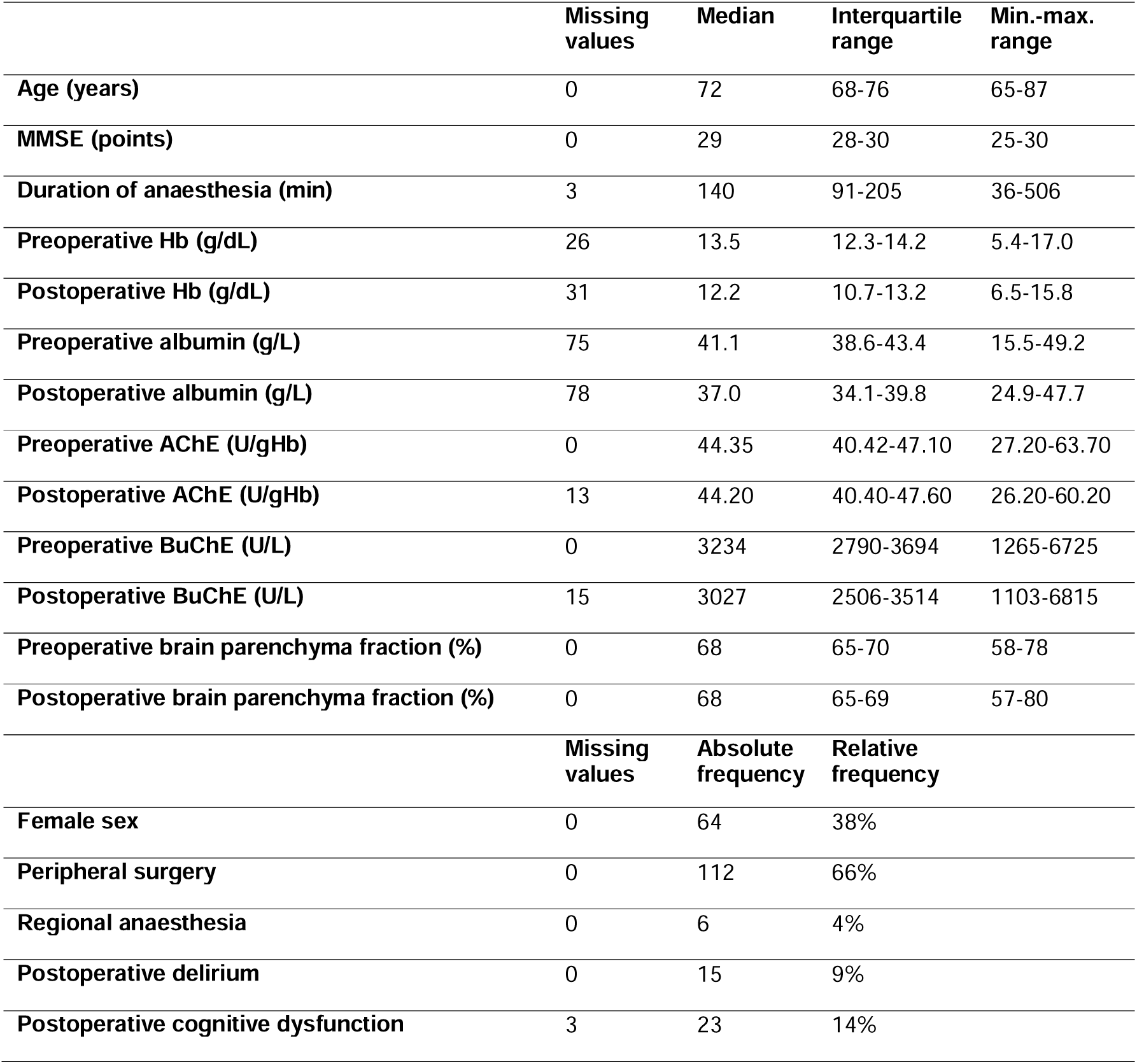
Description of the cohort (N=170 patients)

|  | **Missing values** | **Median** | **Interquartile range** | **Min.-max. range** |
| --- | --- | --- | --- | --- |
| **Age (years)** | 0 | 72 | 68-76 | 65-87 |
| **MMSE (points)** | 0 | 29 | 28-30 | 25-30 |
| **Duration of anaesthesia (min)** | 3 | 140 | 91-205 | 36-506 |
| **Preoperative Hb (g/dL)** | 26 | 13.5 | 12.3-14.2 | 5.4-17.0 |
| **Postoperative Hb (g/dL)** | 31 | 12.2 | 10.7-13.2 | 6.5-15.8 |
| **Preoperative albumin (g/L)** | 75 | 41.1 | 38.6-43.4 | 15.5-49.2 |
| **Postoperative albumin (g/L)** | 78 | 37.0 | 34.1-39.8 | 24.9-47.7 |
| **Preoperative AChE (U/gHb)** | 0 | 44.35 | 40.42-47.10 | 27.20-63.70 |
| **Postoperative AChE (U/gHb)** | 13 | 44.20 | 40.40-47.60 | 26.20-60.20 |
| **Preoperative BuChE (U/L)** | 0 | 3234 | 2790-3694 | 1265-6725 |
| **Postoperative BuChE (U/L)** | 15 | 3027 | 2506-3514 | 1103-6815 |
| **Preoperative brain parenchyma fraction (%)** | 0 | 68 | 65-70 | 58-78 |
| **Postoperative brain parenchyma fraction (%)** | 0 | 68 | 65-69 | 57-80 |
|  | **Missing values** | **Absolute frequency** | **Relative frequency** |  |
| **Female sex** | 0 | 64 | 38% |  |
| **Peripheral surgery** | 0 | 112 | 66% |  |
| **Regional anaesthesia** | 0 | 6 | 4% |  |
| **Postoperative delirium** | 0 | 15 | 9% |  |
| **Postoperative cognitive dysfunction** | 3 | 23 | 14% |  |

There was a strong correlation between NBM volume before surgery and that at follow-up three months after surgery (Pearson’s R=0.96 [0.94; 0.97], p<0.0001, df=168). There was also a strong correlation between ChE activity before surgery and that on the first postoperative day (AChE: Pearson’s R=0.65 [0.55; 0.73], p<0.0001, df=155; BuChE: Pearson’s R=0.68 [0.59; 0.76], p<0.0001, df=153).

The correlation between preoperative AChE and BuChE activity was low (Pearson’s R=0.09 [-0.06; 0.24], p=0.22, df=168), as was the correlation between preoperative ChE activity and NBM volume (AChE: Pearson’s R=0.11 [-0.04; 0.26], p=0.14, df=168; BuChE: Pearson’s R=0.03 [-0.12; 0.18], p=0.7, df=168).

### 3.2 Associations between preoperative ChE activity and NBM atrophy

Neither preoperative AChE activity (B=0.0029 [-0.0057; 0.0116], p=0.5) nor preoperative BuChE activity (B=-1.06 [-2.38; 0.26], p=0.12) was significantly associated with baseline-adjusted postoperative NBM (see Figure 2 and supplementary table S1 for details).

**Figure 2:**
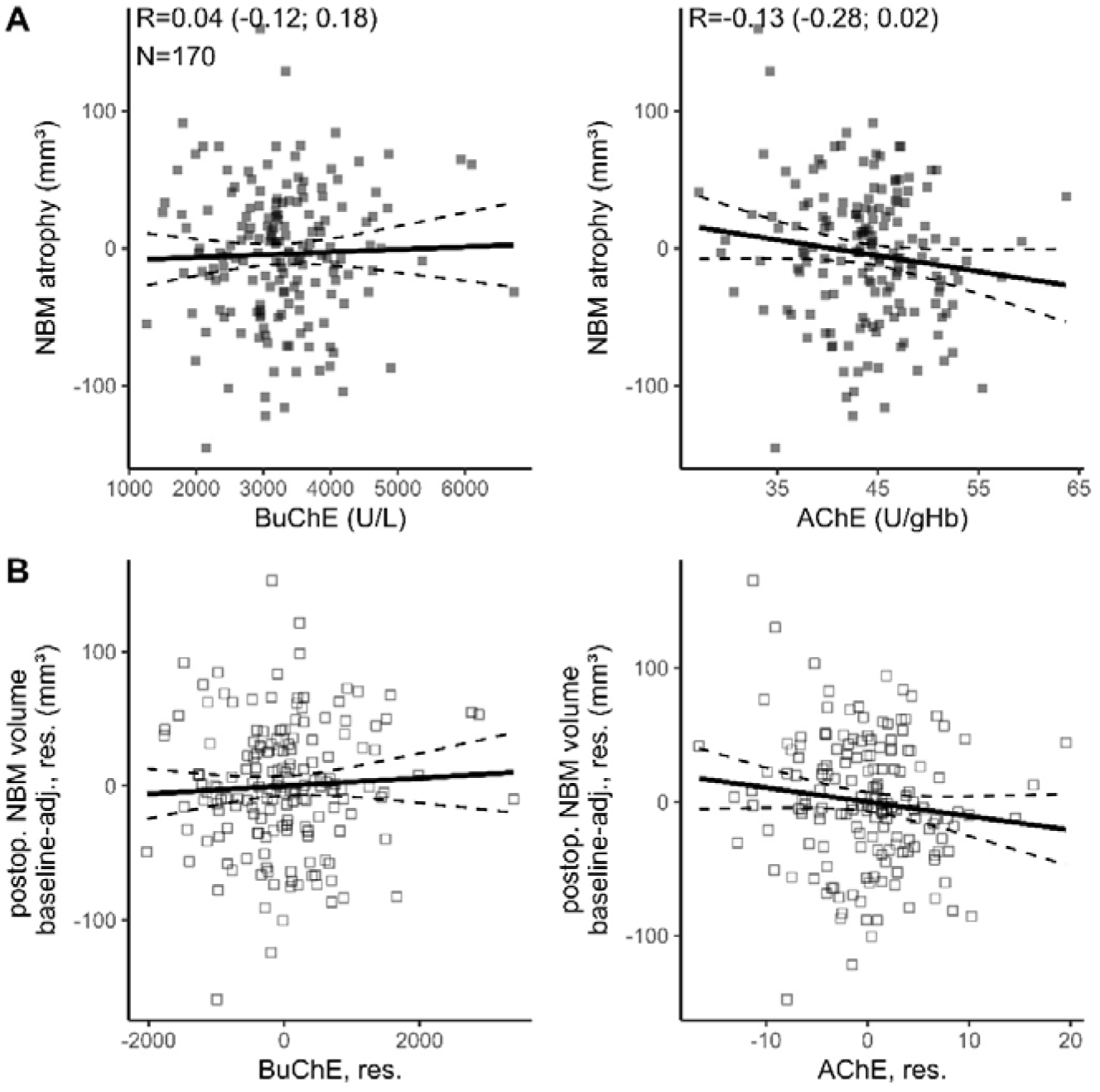
Associations between preoperative ChE activity and postoperative NBM atrophy. Plots with filled squares (_▪_ in A) display native data, e.g. ChE activity on the x-axis and NBM atrophy measured as NBM volume difference (volume_postop._-volume_preop._) on the y-axis, with lower values indicating stronger atrophy. R refers to Pearson’s correlation coefficient (with 95% confidence intervals) between NBM atrophy and ChE activity and N indicates the number of patients. Plots with empty squares (_□_ in B) designate partial regression plots from linear regression models. The y-axis displays residuals (res.) of postoperative NBM volume, after adjustment for baseline volume (baseline-adj.), age, sex and MMSE. The x-axis displays residuals ChE activity, after adjustment for age, sex, MMSE and preoperative NBM volume. Regression lines are given as solid, and dashed lines mark the 95% confidence intervals.

### 3.3 Associations between postoperative ChE activity and NBM atrophy

A lower BuChE activity on the first postoperative day was significantly associated with lower baseline-adjusted postoperative NBM volume, i.e., greater postoperative NBM atrophy (B=0.0141 [0.0055; 0.0227], p=0.0014). Higher postoperative AChE activity was associated with increased postoperative NBM volume, but the association did not reach significance (B=-1.26 [-2.66; 0.14], p=0.077, see Figure 3A+B and supplementary table S2 for details). When postoperative BuChE activity was replaced with preoperative BuChE activity and a postoperative change in BuChE activity, both higher preoperative BuChE activity (B=0.0109 [0.0014; 0.0203], p=0.025) and a greater postoperative increase in BuChE activity (0.0205 [0.0088; 0.0321], p=0.0007) were associated with greater postoperative NBM volume (see Figure 3C+D, and supplementary table S3), but only the latter reached statistical significance after adjustment for multiple testing.

**Figure 3:**
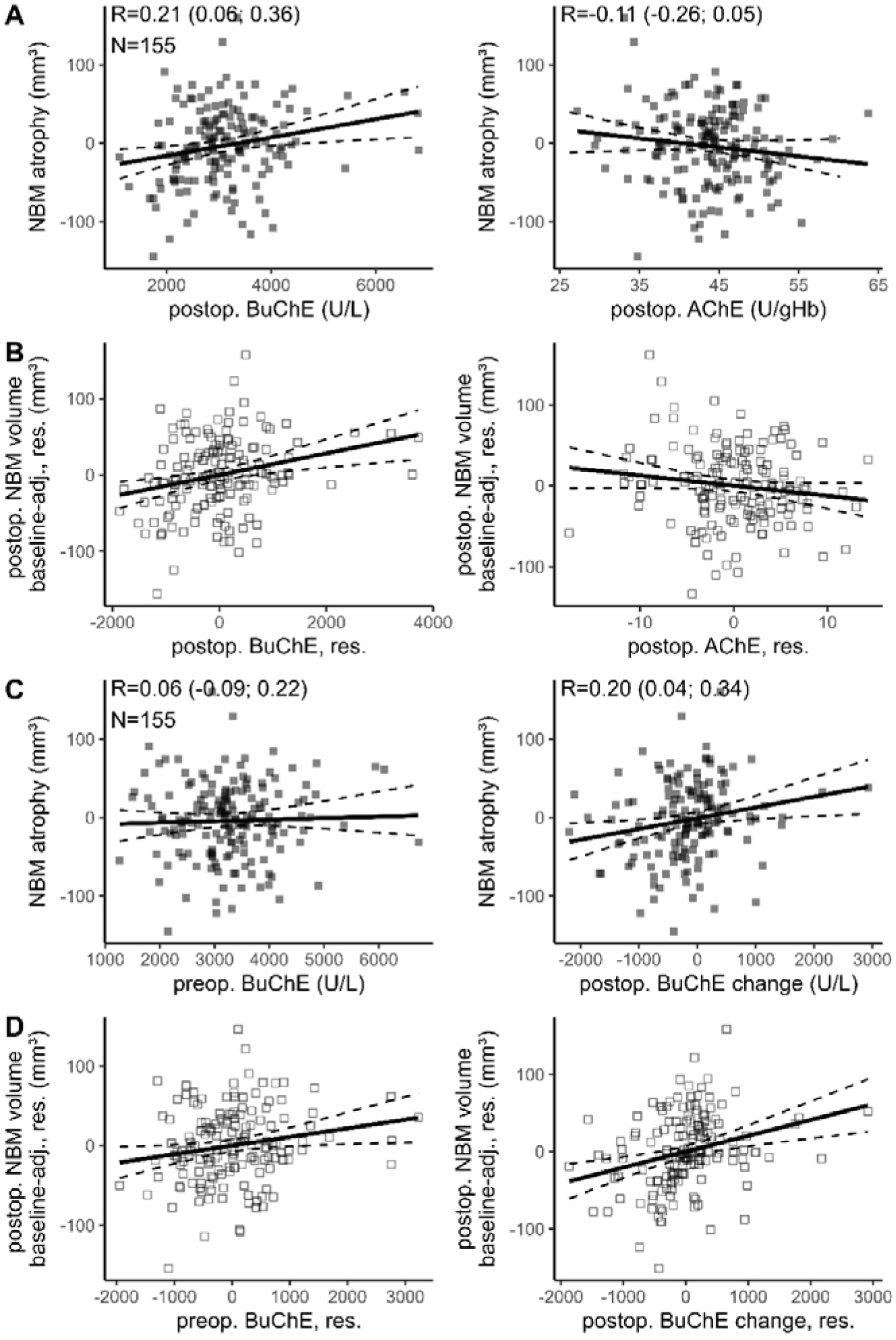
Associations between postoperative ChE activity and NBM atrophy. Plots with filled squares (_▪_ in A and C) display native data, e.g. ChE activity on the x-axis and NBM atrophy measured as NBM volume difference (volumepostop.-volumepreop.) on the y-axis, with lower values indicating stronger atrophy. R refers to Pearson’s correlation coefficient (with 95% confidence intervals) between NBM atrophy and postoperative ChE activity. N indicates the number of patients. Plots with empty squares (_□_ in B and D) designate partial regression plots from linear regression models. The y-axis displays residuals (res.) of postoperative NBM volume, after adjustment for baseline volume (baseline-adj.), age, sex and MMSE. The x-axis displays residuals ChE activity, after adjustment for age, sex, MMSE and preoperative NBM volume. Regression lines are given as solid, and dashed lines mark the 95% confidence intervals. Plots A and B display the results for absolute ChE activity (erythrocyte AChE and free plasma BuChe activity) as measured on the first postoperative day, whereas figures C and D display the associations of preoperative BuChE activity and postoperative change in BuChE activity measured as the difference between postoperative and preoperative BuChE activity (BuChE_postop._-BuChE_preop_). Higher values indicate a postoperative increase in BuChE activity.

### 3.4 Consideration of confounding effects

Since plasma AChE is mostly bound to erythrocytes and BuChE is dependent on liver synthesis capacity, we repeated the analyses with additional adjustment of AChE for Hb values, and adjustment for perioperative albumin levels as a marker of liver function. While there was a trend for higher anaemia-adjusted AChE activity to be associated with NBM atrophy (see supplementary tables S4-S5 and supplementary figure S1), the results for BuChE activity did not change substantially after adjustment for albumin (see supplementary tables S6 and S7). To evaluate the confounding effect of whole-brain atrophy, we replaced NBM volume with brain volume, but no significant associations between postoperative BuChE activity or postoperative changes in BuChE activity and baseline-adjusted brain volume were observed (see supplementary tables S8 and S9).

### 3.5 Associations of POD and POCD with NBM atrophy

Neither POD nor POCD was significantly associated with postoperative NBM atrophy (POD: B=-2.39 [-28.56; 23.79], p=0.9; POCD: B=11.72 [-10.73; 34.18], p=0.30, supplementary tables S10 and S11).

POD did not modulate the association between postoperative BuChE activity (B=-0.004 (-0.894; 1.004, p=0.7, supplementary table S13) or the association between postoperative BuChE activity change and NBM atrophy (B=0.004 [-0.052; 0.059], p=0.9, supplementary table S14). Associations between postoperative ChE activities and NBM atrophy have been stratified by POD for illustration in figure 3.

POCD did not modulate the association between postoperative BuChE activity and NBM atrophy (B=-0.018 [-0.051; 0.015], p=0.29, supplementary table S15, or the association between postoperative BuChE activity change and NBM atrophy (B=0.002 [-0.038; 0.041], p=0.9, supplementary table S16). Associations between postoperative ChE activity and NBM atrophy have been stratified by POCD for illustration in figure 4. In a supplementary analysis, we studied the association between decline in a single cognitive test and NBM atrophy, but no significant associations were observed (supplementary table S12).

**Figure 4:**
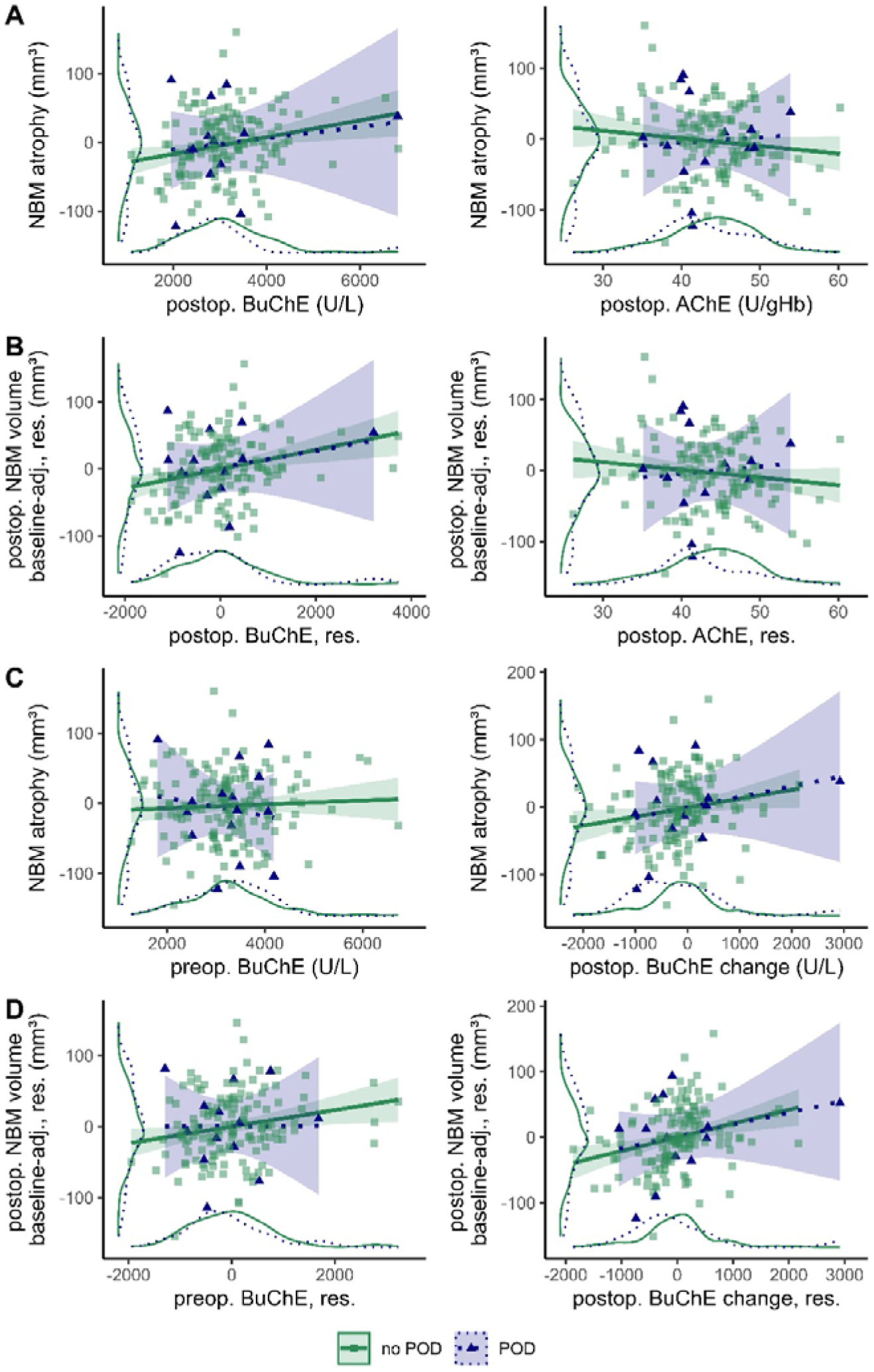
Scatter plots as presented in Figure 3, stratified by occurrence of POD (blue _▴_, dotted lines) and control patients without POD (green _▪_, solid line). Shaded areas around regression lines indicate 95% confidence intervals. Density plots attached to x- and y-axes illustrate the distribution of cholinesterase activity (x-axis) and NBM-atrophy (y-axis) stratified by respective groups.

**Figure 5:**
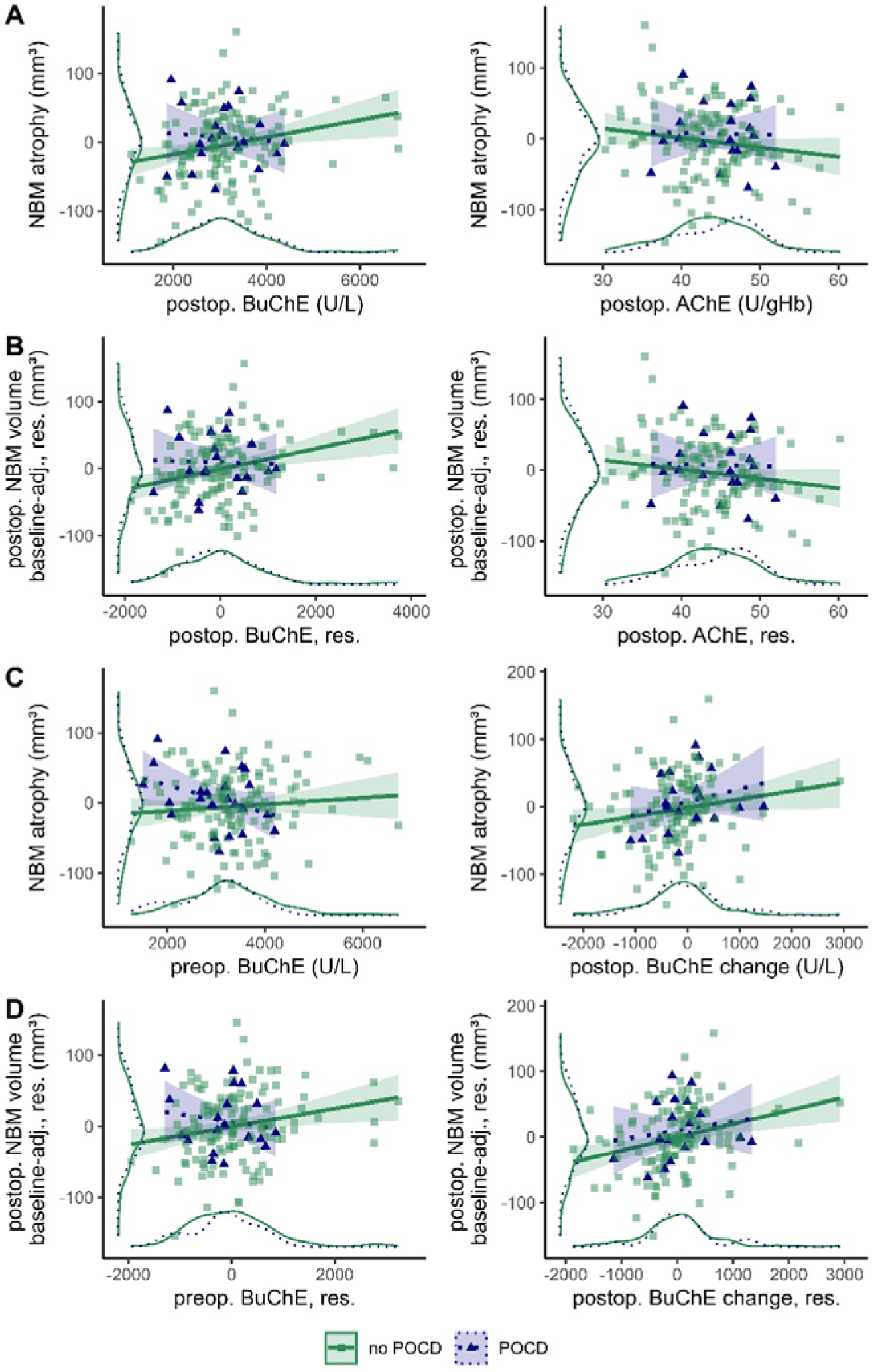
Scatter plots as presented in Figure 3, stratified by occurrence of POD (blue _▴_, dotted lines) and control patients without POCD (green _▪_, solid line). Shaded areas around regression lines indicate 95% confidence intervals. Density plots attached to x- and y-axes illustrate the distribution of cholinesterase activity (x-axis) and NBM-atrophy (y-axis) stratified by respective groups.

## 4 Discussion

Here, we analysed the associations between pre- and postoperative ChE activity and postoperative atrophy of the NBM. We found that lower BuChE activity on the first postoperative day was associated with increased atrophy of the NBM three months after surgery. Postoperative NBM atrophy was not associated with POD or POCD, and neither modulated the association between postoperative BuChE activity and NBM atrophy.

### 4.1 Peripheral BuChE activity is associated with postoperative NBM atrophy

Several lines of evidence indicate that ChE activity is related to neurotoxicity and neurodegeneration. However, the exact molecular mechanism is unknown. Plasma BuChE is synthesised mainly in the liver, but is also present in the brain (Darvesh, Grantham, & Hopkins, 1998). Its relevance for hydrolysation of brain ACh has been described (Cuadra, Summers, & Giacobini, 1994), even though hydrolysation of ACh by BuChE is not its primary physiological function in the brain under normal conditions. Nonetheless, these findings suggest that plasma BuChE activity may also provide information on ChE inhibition and cholinergic neurotransmission in the brain. In fact, it has been suggested that plasma BuChE may act as a detoxifier by binding ChE-inhibitors before they pass the blood-brain barrier, and therefore, low plasma BuChE activity may indicate exhausted detoxification capacity (Darvesh et al., 2003; Johnson & Moore, 2012).

Lower postoperative BuChE activity on the first postoperative day was associated with greater postoperative NBM atrophy, suggesting that brain BuChE inhibition or ACh accumulation can have neurotoxic effects. However, the exact mechanisms are unknown and remain unclear, however, after the BuChE scavenging capacity has been exhausted, neurotoxins, especially substances that inhibit ChE, may be able to exert detrimental effects on the brain.

ACh is synthesised from choline and acetyl-CoA by the choline-acetyltransferase (ChAT). Whereas choline needs to be transported into the neuron or generated from phosphatidylcholine in the neuronal membrane, acetyl-CoA originates from the Krebs-cycle. Both AChE and BuChE are capable of hydrolysing ACh after its release into the synaptic cleft. After hydrolysation, choline is then transported into the neuron and reused for ACh synthesis. Under conditions of ChE inhibition, neurons need to draw on additional acetyl-CoA from the Krebs cycle and choline from the cell membrane for ACh synthesis, with potentially detrimental effects on energy metabolism and cell membrane integrity. Notably, cholinergic neurons constantly release low amounts of ACh, aggravating the need for a constant supply of acetyl-CoA and choline as well as a stable energy supply (Siegel, 2006).

In fact, neurodegeneration after exposure to ChE inhibitors has been reported in the past literature. For example, poisoning, but not therapy, with specific ChE-inhibitors (organophosphates and carbamates) has been reported to increase the risk for Parkinson dementia (Chuang et al., 2017; Wang et al., 2014; Yan et al., 2018) and cortical atrophy occurs in veterans exposed to sarin during the Gulf War (Chao et al., 2011; Chao et al., 2010; Krengel et al., 2024). With respect to Parkinson’s disease, NBM atrophy is regularly reported in patients with Lewy-body-related diseases (Conti et al., 2024; Pereira et al., 2020; Rogozinski et al., 2022), indicating a particular vulnerability of cholinergic neurons, in accordance with our findings. For Gulf War illness, atrophy of the cholinergic system is less consistently reported, but past reports point to possible effects on cholinergic brainstem nuclei (Christova et al., 2017; Zhang et al., 2020), white matter tracts originating from the NBM, e.g., in the temporal stem(Yamasue et al., 2007) and cortical atrophy in close proximity to the NBM (Chao et al., 2010).

ChE-inhibitors, such as carbamates and organophosphorus compounds, are still commonly used as pesticides and may contribute to the global burden of neurodegenerative diseases in frequently exposed populations, such as those in agricultural professions (Bustamante et al., 2025; Macheka et al., 2024). Even though persistence in ground water cannot be fully ruled out, it seems unlikely that environmental exposure to these agents accounts for postoperative BuChE inhibition and NBM atrophy in our cohort. Modern anaesthetic agents are generally not regarded as directly inhibiting BuChE to a clinically relevant degree. However, notable indirect interactions between BuChE and certain anaesthetics, with respect to muscle relaxants as well as individual variability in enzyme activity have been described. Indeed, BuChE is a pleiotropic enzyme involved in the metabolism of various drugs, many of which are regularly used in perioperative medicine, e.g., succinylcholine, mivacurium, acetylsalicylic acid, procaine, physostigmine or neostigmine (Masson, Shaihutdinova, & Lockridge, 2023). On the other hand, metoclopramide (Kao, Tellez, & Turner, 1990), local anesthetics (Davie, 1977), non-depolarising muscle relaxants (Mirakhur, Ferres, & Lavery, 1983), cytostatics (Wood, 1991), topically applied AChE inhibitors (Eilderton, Farmati, & Zsigmond, 1968), coumarin derivatives (Macklin & Schwans, 2020), and preoperative alimentation have been found to have systemic inhibitory effects on BuChE activity (Bestas, Goksu, & Erhan, 2013; McGehee et al., 2000), and several other drugs commonly used during anaesthesia are also suspected to affect BuChE activity (Andersson, Moller, & Wildgaard, 2019). However, little is known about whether these drugs act as inhibitors or competitive substrates at BuChE. On the other hand, there is indirect evidence that the commonly used anaesthetic sevoflurane can significantly impact BuChE activity: It has been shown that plasma fluoride concentrations of 35µmol/L occur after sevoflurane anaesthesia (Sondekoppam et al., 2020). These concentrations have been reported to inhibit BuChE activity by almost 50% with only negligible effects on AChE activity (Kambam, Parris, Naukam, Franks, & Sastry, 1990). While sevoflurane exposure was not recorded in this study and the impact of sevoflurane on BuChE activity may therefore only act as an example from the literature, it seems possible that perioperative drug administration and unspecific BuChE inhibition account for impaired enzyme activity and the toxic effects of ACh accumulation in the brain.

In addition to BuChE inhibition by ChE-inhibitors, decreased BuChE expression may account for lower BuChE activity, implying that other nonenzymatic actions of BuChE may account for its association with postoperative NBM volume (Pope & Brimijoin, 2018). Some studies reported that reduced BuChE activity after cardiopulmonal bypass was mainly, but not fully, explained by dilution and decreased BuChe concentration (Rump et al., 1999). Since the liver is the main source of plasma BuChE, we considered liver synthesis capacity or postoperative plasma dilution a major confounding factor. However, our results did not change substantially after adjusting analyses for albumin as a surrogate parameter for liver function and haemodilution. However, the BuChE plasma concentration was not measured in this study, and little is known about the regulation of BuChE expression. Hence, the mechanisms by which lower BuChE expression could affect postoperative NBM volume remain hypothetical (Johnson & Moore, 2012). In addition, there is a well-known genetic influence on BuChE activity, that was not addressed in this study (Johnson & Moore, 2012). However, in the case of a genetic predisposition, we would have expected associations with preoperative BuChE activity rather than with postoperative values.

### 4.2 No evidence for NBM atrophy in patients with POD or POCD

In a previous study, we reported lower postoperative BuChE activity in patients with POD(Muller et al., 2019). We hence hypothesised that postoperative BuChE activity may mediate or moderate NBM atrophy in patients with POD or POCD. However, there was no group difference in the baseline-adjusted postoperative NBM volume between patients with POD or POCD and the respective control patients, and neither POD nor POCD modulated the association between postoperative BuChE activity and NBM atrophy. In fact, in another subsample from the BioCog study including only patients who underwent abdominal surgery, BuChE activity was lower in patients with POD, but did not achieve significance (Bosancic et al., 2022).

However, there are several concerns regarding this analysis. First, the percentage of patients lot to follow-up was as high as 34% of patients who underwent MRI before surgery. Notably, the POD rate in the sample presented here was 9%, which is considerably lower than the rate reported for the whole cohort (Florian Lammers-Lietz et al., 2024). This problem is inherent to the study design, as especially frail patients at high risk for POD may be more likely to refuse participation in exhausting procedures such as MRI, and patients with postoperative complications need more time to recover and may be too weak to participate in follow-up assessments. Hence, this study may suffer from selection bias because only patients with favourable postoperative outcomes were analysed. In fact, previous studies have questioned the association of short episodes of POD with POCD (Franck et al., 2016) and postoperative neuronal damage is more prominent in severe POD (Cavallari et al., 2017). On the other hand, postoperative alterations in the basal forebrain cholinergic system have been reported to be associated with postoperative cognition, and POD patients were found to have increased global brain atrophy (M. Heinrich et al., 2024a; Kant et al., 2022; F. Lammers-Lietz et al., 2024). In addition, the period in which POD may impact cognitive trajectories may be much longer, i.e., spanning several years (Racine et al., 2020; Teipel et al., 2018). Hence, longer follow-up periods may be necessary to substantiate associations between POD, postoperative cognitive function and BuChE-mediated NBM atrophy.

### 4.3 Limitations

In this study, POCD was assessed in accordance with criteria proposed at the time of study initiation and reflects a postoperative decline in performance in a neuropsychological testing battery compared with a nonsurgical reference group (Borchers et al., 2021; Feinkohl et al., 2020; Rasmussen et al., 2001). However, the nomenclature of postoperative neurocognitive disorders (NCD) was modified and requires an additional subjective cognitive concern (minor postoperative NCD) and eventually impaired functioning in activities of daily living (major NCD).

As outlined above, sevoflurane anaesthesia may be a major determinant of postoperative BuChE activity, which was not recorded in the BioCog study. In addition, BuChE protein expression was not measured in this study. Hence, it is unknown whether BuChE activity results from impaired BuChE synthesis or from enzyme inhibition. In a proteome analysis of a subsample of matched cases from the BioCog study, a more complex association between BuChE abundance and POD was reported (Lamping et al., 2025). However, the number of patients in the sample described here and the sample with proteome data was too low for a thorough analysis.

We observed two interesting associations that were not statistically significant. In the supplementary analyses, higher preoperative anaemia-adjusted AChE activity was associated with lower baseline-adjusted postoperative NBM volume (table S4, figure S1), and lower preoperative BuChE was also associated with lower postoperative NBM volume after controlling for postoperative changes in BuChE activity (table S3). The significance of these findings may be proven in studies with greater statistical power, but discussing the biomedical relevance of these findings is beyond the scope of this work.

### 4.4 Conclusion

This is the first report on associations between peripheral cholinesterase activity and atrophy in the central nervous cholinergic system. Cholinesterases play a central role in mediating neuronal, endocrine and immune functions, and atrophy of the cholinergic system occurs in various conditions, including the most common neurodegenerative diseases. Our results will contribute to the overall understanding of pathomechanisms under these conditions. However, we emphasize the urgent need to understand the molecular pathways that link cholinesterase activity with the atrophy of cholinergic nuclei, before these pathways can be considered as targets for potential new therapies.

## 5 Statements and declarations

### 5.2 Author contributions

Florian Lammers-Lietz: Conceptualisation, formal analysis, investigation, methodology, visualization, writing – original draft. Maria Heinrich: Conceptualization, investigation. Kai Kappert: writing – review and editing. Georg Winterer: Project administration, funding acquisition, resources, methodology. Claudia Spies: Project administration, funding acquisition, resources, methodology. Anika Müller: Conceptualization, investigation, methodology, supervision.

### 5.3 Ethics approval

All procedures were approved by the local ethics committees in Berlin, Germany (Charité-Universitätsmedizin Berlin, EA2/092/14) and Utrecht, Netherlands (University Medical Center Utrecht, 14-469) and conducted in line with the Declaration of Helsinki.

### 5.4 Consent to participate

All the participants provided written informed consent prior to inclusion.

### 5.5 Sex and gender

Data on the patients’ gender have not been collected, and we refer to “sex” throughout the manuscript.

### 5.6 Data availability

Participant data may be made available upon request following publication to researchers who provide a methodologically sound proposal in accordance with applicable legal and regulatory restrictions after careful review of each individual request. Access will only be granted in cases where the potential receiver of the data and purpose of the analysis is covered by the patients’ informed consent and applicable legal regulations.

Proposals for data analysis must be directed to both. Analyses will be limited to those approved in appropriate ethics and governance arrangements. All study documents that do not identify individuals (e.g. study protocol, informed consent form template) will be freely available on request.

### 5.7 Use of artificial intelligence and assistance in scientific writing

Neither artificial intelligence tools nor a professional medical writer has assisted in manuscript preparation.

