## supplement for "Associations between blood cholinesterase activity and postoperative basal forebrain cholinergic system atrophy: Results from the BioCog cohort study"

**Table S1: Details for a linear regression model of baseline-adjusted postoperative NBM volume (mm³) with preoperative ChE activities as independent variables of interest**

| | B (95% confidence intervale), p-value |
|---|---|
| Intercept* | 1742.6078 (1729.0495; 1756.1661), p=0.0000 |
| Preoperative NBM volume (mm²) | 0.9529 (0.9016; 1.0042), p=0.0000 |
| Age (years) | -0.0958 (-1.6553; 1.4636), p=0.9036 |
| MMSE (points) | 4.1524 (-2.0976; 10.4024), p=0.1914 |
| Female sex | -1.3082 (-18.3352; 15.7188), p=0.8796 |
| Preoperative BuChE activity (U/L) | 0.0029 (-0.0057; 0.0116), p=0.5018 |
| Preoperative AChE activity (U/gHb) | -1.0576 (-2.3759; 0.2608), p=0.1151 |
| N | 170 |
| $R^2$ | 0.9186 |

* Intercept for: Mean preoperative NBM volume, age of 75 years, MMSE of 29p, male sex, mean ChE activity

**Table S2: Details for a linear regression model of baseline-adjusted postoperative NBM volume (mm³) with postoperative ChE activities as independent variables of interest**

| | B (95% confidence intervale), p-value |
|---|---|
| Intercept* | 1742.3418 (1728.4839; 1756.1997), p=0.0000 |
| Preoperative NBM volume (mm²) | 0.9514 (0.8972; 1.0056), p=0.0000 |
| Age (years) | 0.3922 (-1.2398; 2.0242), p=0.6356 |
| MMSE (points) | 5.2323 (-1.4948; 11.9594), p=0.1264 |
| Female sex | 2.6581 (-14.7871; 20.1033), p=0.7638 |
| Postoperative BuChE activity (U/L) | 0.0141 (0.0055; 0.0227), p=0.0014 |
| Postoperative AChE activity (U/gHb) | -1.2623 (-2.6640; 0.1393), p=0.0772 |
| N | 155 |
| $R^2$ | 0.9165 |

* Intercept for: Mean preoperative NBM volume, age of 75 years, MMSE of 29p, male sex, mean ChE activity

**Table S3: Details for a linear regression model of baseline-adjusted postoperative NBM volume (mm³). including preoperative BuChE activity and postoperative BuChE activity change**

| | B (95% confidence intervale), p-value |
|---|---|
| Intercept* | 1742.1766 (1728.3862; 1755.9669), p=0.0000 |
| Preoperative NBM volume (mm²) | 0.9514 (0.8975; 1.0054), p=0.0000 |
| Age (years) | 0.4050 (-1.2189; 2.0288), p=0.6229 |
| MMSE (points) | 5.4400 (-1.2582; 12.1383), p=0.1106 |
| Female sex | 2.9954 (-14.3670; 20.3577), p=0.7336 |
| Preoperative BuChE activity (U/L) | 0.0109 (0.0014; 0.0203), p=0.0248 |
| Postoperative BuChE change (U/L) | 0.0205 (0.0088; 0.0321), p=0.0007 |
| Postoperative AChE activity (U/gHb) | -1.3299 (-2.7271; 0.0672), p=0.0619 |
| N | 155 |
| $R^2$ | 0.9179 |

* Intercept for: Mean preoperative NBM volume, age of 75 years, MMSE of 29p, male sex, mean ChE activity

To adjust for a possible confounding effect of anaemia, we repeated the analysis with AChE activity measured in [U/dL], referred to as $AChE_{vol}$, by calculating the product of AChE activity [in U/gHb] with haemoglobin [in g/dL] before surgery.

When $AChE_{vol}$ was included in the model, there was a significant association between preoperative $AChE_{vol}$ activity and baseline-adjusted postoperative NBM volume (B=-0.0837 [-0.1562; -0.0113], p=0.024, see Figure S 1 and supplementary table S4 for details). These findings suggest that higher preoperative AChE activity is associated with greater postoperative NBM atrophy.

For ChE activity measured on the first postoperative day, the results remained unaltered after replacing postoperative AChE activity with $AChE_{vol}$ activity (supplementary table S5).

**Table S4: Details for a linear regression model of baseline-adjusted postoperative NBM volume ($mm^3$). AChE activity is measured in [U/dL] rather than [U/gHb].**

| | B (95% confidence intervale), p-value |
|---|---|
| Intercept* | 1743.7489 (1729.8964; 1757.6014), p=0.0000 |
| Preoperative NBM volume ($mm^2$) | 0.9476 (0.8940; 1.0011), p=0.0000 |
| Age (years) | 0.2347 (-1.3196; 1.7889), p=0.7657 |
| MMSE (points) | 4.0921 (-2.4348; 10.6190), p=0.2172 |
| Female sex | -0.1722 (-17.7658; 17.4215), p=0.9846 |
| Preoperative BuChE activity (U/L) | 0.0045 (-0.0044; 0.0134), p=0.3221 |
| Preoperative AChE activity ($AChE_{vol}$, U/dL)** | -0.0837 (-0.1562; -0.0113), p=0.0238 |
| N*** | 144 |
| $R^2$ | 0.9208 |

* Intercept for: Mean preoperative NBM volume, age of 75 years, MMSE of 29p, male sex, mean ChE activity

** Value has been calculated as the product of preoperative AChE activity in [U/gHb] and preoperative Hb level in [g/dL]

*** lower samples size arises from missing values in preoperative Hb levels

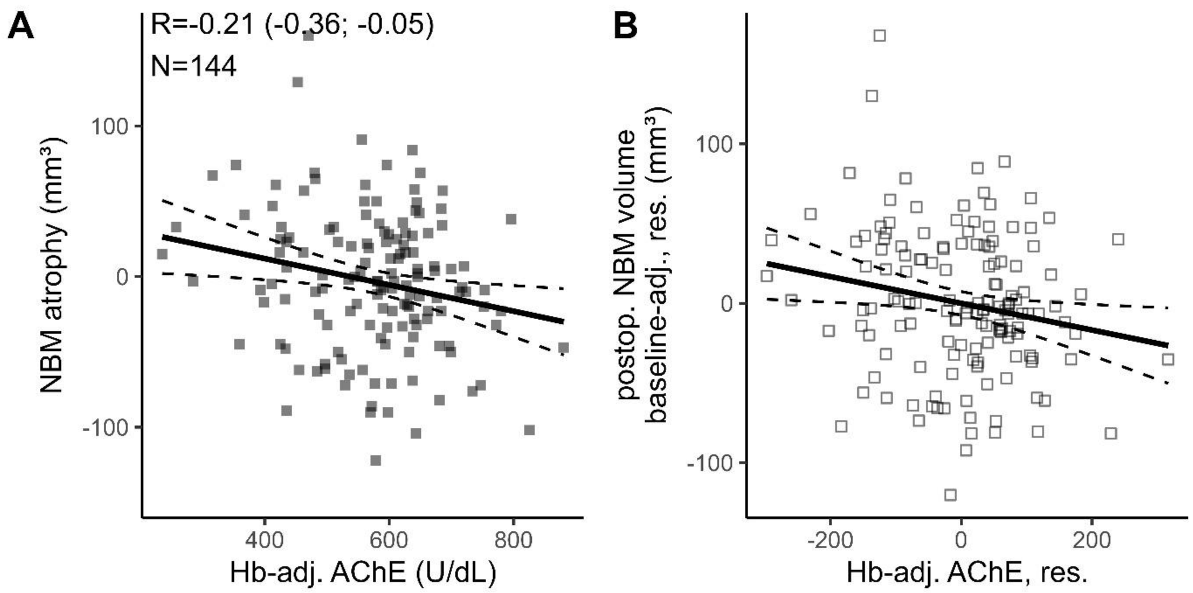


Figure S 1: Associations between preoperative hemoglobin (Hb)-adjusted AChE activity and postoperative NBM atrophy, illustrating findings from supplementary table S2. Plots with filled squares (■ in A) display native data, e.g. ChE activity on the x-axis and NBM atrophy measured as NBM volume difference ($volume_{postop.}$-$volume_{preop.}$) on the y-axis, with lower values indicating stronger atrophy. R refers to Pearson's correlation coefficient (with 95% confidence intervals) between NBM atrophy and ChE activity and N indicates the number of patients. Plots with empty squares (□ in B) designate partial regression plots from linear regression models. The y-axis displays residuals (res.) of postoperative NBM volume, after adjustment for baseline volume (baseline-adj.), age, sex and MMSE. The x-axis display residuals ChE activity, after adjustment for age, sex, MMSE and preoperative NBM volume. Regression lines are given as solid, and dashed lines mark the 95% confidence intervals.

**Table S5: Details for a linear regression model of baseline-adjusted postoperative NBM volume (mm³). AChE activity is measured in [U/dL] rather than [U/gHb].**

| | B (95% confidence intervale), p-value |
|---|---|
| Intercept* | 1745.5787 (1731.1479; 1760.0094), p=0.0000 |
| Preoperative NBM volume (mm²) | 0.9477 (0.8886; 1.0067), p=0.0000 |
| Age (years) | 0.5672 (-1.0891; 2.2234), p=0.4992 |
| MMSE (points) | 3.2921 (-3.9379; 10.5220), p=0.3692 |
| Female sex | -0.8120 (-19.2754; 17.6514), p=0.9308 |
| Postoperative BuChE activity (U/L) | 0.0126 (0.0039; 0.0212), p=0.0047 |
| Postoperative AChE activity ($AChE_{vol}$, U/dL)** | -0.0351 (-0.1129; 0.0428), p=0.3745 |
| N*** | 132 |
| $R^2$ | 0.9125 |

* Intercept for: Mean preoperative NBM volume, age of 75 years, MMSE of 29p, male sex, mean ChE activity

** Value has been calculated as the product of preoperative AChE activity in [U/gHb] and preoperative Hb level in [g/dL]

*** lower sample size arises from missing values in postoperative Hb levels

BuChE activity correlated with albumin level before surgery (Pearson’s R=0.31 [0.11; 0.48], p=0.003, N=95) and on the first postoperative day (Pearson’s R=0.35 [0.15; 0.52], p=0.001, N=85). The perioperative change in BuChE activity was correlated with the perioperative change in albumin level (Pearson’s R=0.48 [0.29; 0.62], p<0.001, N=85).

**Table S6: Details for a linear regression model of baseline-adjusted postoperative NBM volume ($mm^3$) with additional adjustment for postoperative albumin levels**

| | **B (95% confidence intervale), p-value** |
|---|---|
| Intercept* | 1741.0834 (1723.0632; 1759.1035), p=0.0000 |
| Preoperative NBM volume ($mm^2$) | 0.9287 (0.8591; 0.9982), p=0.0000 |
| Age (years) | 0.5215 (-1.4895; 2.5325), p=0.6071 |
| MMSE (points) | -0.8450 (-9.6573; 7.9672), p=0.8491 |
| Female sex | -2.5049 (-25.3250; 20.3151), p=0.8276 |
| Postoperative BuChE activity (U/L) | 0.0110 (0.0008; 0.0213), p=0.0355 |
| Postoperative AChE activity (U/gHb) | -1.3147 (-2.9859; 0.3566), p=0.1214 |
| Postoperative albumin levels (g/L) | 2.1303 (-0.1363; 4.3970), p=0.0651 |
| N** | 86 |
| $R^2$ | 0.9195 |

* Intercept for: Mean preoperative NBM volume, age of 75 years, MMSE of 29p, male sex, mean ChE activity and mean albumin levels

** lower sample size arises from missing values in pre- and postoperative albumin levels

**Table S7: Details for a linear regression model of baseline-adjusted postoperative NBM volume (mm³). including preoperative BuChE activity and postoperative BuChE activity change with additional adjustment for postoperative albumin levels**

| | B (95% confidence intervale), p-value |
|---|---|
| Intercept* | 1739.1243 (1720.8270; 1757.4215), p=0.0000 |
| Preoperative NBM volume (mm²) | 0.9264 (0.8570; 0.9958), p=0.0000 |
| Age (years) | 0.6723 (-1.3602; 2.7048), p=0.5120 |
| MMSE (points) | -1.3091 (-10.1413; 7.5232), p=0.7686 |
| Female sex | 1.7441 (-21.7729; 25.2612), p=0.8829 |
| Preoperative BuChE activity (U/L) | 0.0074 (-0.0040; 0.0189), p=0.2007 |
| Postoperative BuChE change (U/L) | 0.0172 (0.0016; 0.0329), p=0.0314 |
| Postoperative AChE activity (U/gHb) | -1.4037 (-3.0840; 0.2767), p=0.1003 |
| Preoperative albumin level (g/L) | 2.6765 (-0.0197; 5.3727), p=0.0517 |
| Postoperative albumin change (g/L) | 1.4109 (-1.1729; 3.9946), p=0.2802 |
| N** | 85 |
| $R^2$ | 0.9234 |

* Intercept for: Mean preoperative NBM volume, age of 75 years, MMSE of 29p, male sex, mean ChE activity and mean albumin levels

** lower sample size arises from missing values in postoperative albumin levels

**Table S8: Details for a linear regression model of baseline-adjusted postoperative brain volume (cm³) with postoperative ChE activities as independent variables of interest**

| | B (95% confidence intervale), p-value |
|---|---|
| Intercept* | 994.876 (988.810; 1000.942), p=0.0000 |
| Preoperative brain volume (cm²) | 0.9823 (0.9418; 1.0227), p=0.0000 |
| Age (years) | -151.2096 (-885.3049; 582.8857), p=0.6846 |
| MMSE (points) | 340.7295 (-2528.2298; 3209.6888), p=0.8148 |
| Female sex | -1107.5221 (-8639.5128; 6424.4686), p=0.7718 |
| Postoperative BuChE activity (U/L) | 2.2275 (-1.4575; 5.9124), p=0.2342 |
| Postoperative AChE activity (U/gHb) | 20.8736 (-578.0610; 619.8083), p=0.9452 |
| N | 155 |
| $R^2$ | 0.9589 |

* Intercept for: Mean preoperative brain volume, age of 75 years, MMSE of 29p, male sex, mean ChE activity

**Table S9: Details for a linear regression model of baseline-adjusted postoperative brain volume ($cm^3$). including preoperative BuChE activity and postoperative BuChE activity change**

| | B (95% confidence intervale), p-value |
|---|---|
| Intercept* | 994.862 (988.781; 1000.943), p=0.0000 |
| Preoperative brain volume ($mm^2$) | 0.9824 (0.9419; 1.0230), p=0.0000 |
| Age (years) | -147.9915 (-883.8730; 587.8900), p=0.6916 |
| MMSE (points) | 371.6071 (-2505.9588; 3249.1731), p=0.7989 |
| Female sex | -1068.6603 (-8619.3053; 6481.9848), p=0.7801 |
| Preoperative BuChE activity (U/L) | 1.7133 (-2.3796; 5.8062), p=0.4094 |
| Postoperative BuChE change (U/L) | 3.2198 (-1.8027; 8.2423), p=0.2072 |
| Postoperative AChE activity (U/gHb) | 10.3713 (-591.0328; 611.7753), p=0.9729 |
| N | 155 |
| $R^2$ | 0.9590 |

* Intercept for: Mean preoperative brain volume, age of 75 years, MMSE of 29p, male sex, mean ChE activity

**Table S10: Details for a linear regression model of associations of POD with baseline-adjusted postoperative NBM volume (mm³)**

| | B (95% confidence intervale), p-value |
|---|---|
| Intercept* | 1742.8823 (1729.0418; 1756.7228), p=0.0000 |
| Preoperative NBM volume (mm²) | 0.9499 (0.8985; 1.0012), p=0.0000 |
| Age (years) | 0.0438 (-1.5125; 1.6001), p=0.9557 |
| MMSE (points) | 4.0752 (-2.2448; 10.3952), p=0.2047 |
| Female sex | -0.8540 (-17.9433; 16.2353), p=0.9215 |
| POD** | -2.3852 (-28.5643; 23.7939), p=0.8574 |
| N | 170 |
| $R^2$ | 0.9172 |

* Intercept for: Mean preoperative NBM volume, age of 75 years, MMSE of 29p, male sex, no POD

** POD, postoperative delirium

**Table S11: Details for a linear regression model of associations of POCD with baseline-adjusted postoperative NBM volume (mm³)**

| | B (95% confidence intervale), p-value |
|---|---|
| Intercept* | 1740.3347 (1726.0596; 1754.6098), p=0.0000 |
| Preoperative NBM volume (mm²) | 0.9487 (0.8975; 0.9999), p=0.0000 |
| Age (years) | -0.0725 (-1.6929; 1.5480), p=0.9297 |
| MMSE (points) | 5.1162 (-1.2185; 11.4508), p=0.1127 |
| Female sex | 0.8516 (-16.2776; 17.9807), p=0.9219 |
| POCD** after three months | 11.7246 (-10.7267; 34.1759), p=0.3040 |
| N*** | 167 |
| $R^2$ | 0.9189 |

* Intercept for: Mean preoperative NBM volume, age of 75 years, MMSE of 29p, male sex, no POCD

** POCD, postoperative cognitive dysfunction

*** lower N arises from patients without POCD assessment after three months

**Table S12: Associations of longitudinal changes in cognitive test performance with NBM atrophy.**

Model specifications correspond to the model described in supplementary table S11, but instead of POCD as a variable, a cognitive decline in a specific cognitive has been included in the model. For brevity, only the regression coefficient (B) for the cognitive decline variable is given.

Performance decline in each score has been transformed into a z-score according to performance decline in a nonsurgical reference group and dichotomized at a cut-off value of z<-1.96, indicating a cognitive decline more extreme than 95% of the reference group for a normally distributed test parameter.

| Cognitive test parameter* | N with z<-1.96** | B (95% CI)*** | p-value |
|---|---|---|---|
| **PAL (first trial memory score)** | 6 | 24.03 (-16.39; 64.45) | 0.24 |
| **VRM (immediate recall)** | 3 | -19.54 (-76.10; 37.01) | 0.5 |
| **VRM (delayed recognition)** | 19 | 8.76 (-15.07; 32.59) | 0.47 |
| **SSP (span length)** | 1 | 52.17 (-45.35; 149.69) | 0.29 |
| **GPT (time)** | 16 | 7.36 (-18.79; 33.51) | 0.6 |
| **SRT (latency for correct responses)** | 4 | 15.53 (-34.45; 65.52) | 0.5 |
| **TMT-B (completion time)** | 9 | 5.23 (-27.87: 38.32) | 0.8 |

* For each cognitive test, a separate regression analysis has been conducted

** Total sample size in N=170 for all tests. Missing values have been imputed as described in the manuscript.

*** Regression coefficients (B) have been adjusted for age, sex and baseline MMSE score. The regression coefficient can be interpreted as the mean difference in postoperative NBM volume between patients with a z-score below and above -1.96. Negative volumes indicate lower volumes in the group with cognitive decline.

**Table S13: Details for a linear regression model of a modulator effect (interaction) of POD on the association between postoperative BuChE and baseline-adjusted postoperative NBM volume ($mm^3$)**

| | B (95% confidence intervale), p-value |
|---|---|
| Intercept* | 1744.1082 (1726.0030; 1762.2134), p=0.0000 |
| Preoperative NBM volume ($mm^2$) | 0.9489 (0.8939; 1.0039), p=0.0000 |
| Age (years) | 0.3836 (-1.2735; 2.0408), p=0.6480 |
| MMSE (points) | 4.7600 (-2.0692; 11.5893), p=0.1705 |
| Female sex | 1.8015 (-15.8538; 19.4569), p=0.8405 |
| Postoperative BuChE activity (U/L) | 0.0113 (-0.0011; 0.0237), p=0.0735 |
| POD** | 3.2080 (-24.6721; 31.0881), p=0.8204 |
| Interaction between BuChE activity and POD** | -0.0045 (-0.0292; 0.0202), p=0.7200 |
| N | 155 |
| $R^2$ | 0.9148 |

* Intercept for: Mean preoperative NBM volume, age of 75 years, MMSE of 29p, male sex, no POD

** POD, postoperative delirium

**Table S14: Details for a linear regression model of a modulator effect (interaction) of POD on the association between postoperative BuChE activity change and baseline-adjusted postoperative NBM volume (mm³)**

| | B (95% confidence intervale), p-value |
|---|---|
| Intercept* | 1740.3535 (1720.7813; 1759.9257), p=0.0000 |
| Preoperative NBM volume (mm²) | 0.9546 (0.8991; 1.0101), p=0.0000 |
| Age (years) | 0.3291 (-1.3434; 2.0017), p=0.6979 |
| MMSE (points) | 4.4687 (-2.4754; 11.4129), p=0.2055 |
| Female sex | 0.6998 (-17.2452; 18.6448), p=0.9387 |
| Postoperative BuChE change (U/L) | 0.0161 (-0.0116; 0.0438), p=0.2529 |
| POD** | 2.0545 (-28.3450; 32.4540), p=0.8939 |
| Interaction between postoperative BuChE change (U/L) and POD | 0.0037 (-0.0519; 0.0593), p=0.8954 |
| N*** | 154 |
| $R^2$ | 0.9136 |

* Intercept for: Mean preoperative NBM volume, age of 75 years, MMSE of 29p, male sex, no POD

** POD, postoperative delirium

*** One potentially influential outlier has been excluded

**Table S15: Details for a linear regression model of a modulator effect (interaction) of POCD on the association between postoperative BuChE and baseline-adjusted postoperative NBM volume (mm³)**

| | **B (95% confidence intervale), p-value** |
|---|---|
| Intercept* | 1746.2371 (1730.1800; 1762.2941), p=0.0000 |
| Preoperative NBM volume (mm²) | 0.9512 (0.8965; 1.0060), p=0.0000 |
| Age (years) | 0.3059 (-1.4016; 2.0133), p=0.7238 |
| MMSE (points) | 5.7300 (-1.1021; 12.5621), p=0.0996 |
| Female sex | 2.7296 (-14.8906; 20.3498), p=0.7599 |
| Postoperative BuChE activity (U/L) | 0.0049 (-0.0116; 0.0213), p=0.5578 |
| POCD** | 9.5425 (-14.8734; 33.9583), p=0.4411 |
| Interaction between BuChE activity and POCD** | -0.0178 (-0.0506; 0.0151), p=0.2872 |
| N*** | 153 |
| $R^2$ | 0.9165 |

* Intercept for: Mean preoperative NBM volume, age of 75 years, MMSE of 29p, male sex, no POCD

** POCD, postoperative cognitive dysfunction after three months

*** two patients have missing values for POCD assessment

**Table S16: Details for a linear regression model of a modulator effect (interaction) of POCD on the association between postoperative BuChE activity change and baseline-adjusted postoperative NBM volume (mm³)**

| | B (95% confidence intervale), p-value |
|---|---|
| Intercept* | 1745.8108 (1729.2777; 1762.3439), p=0.0000 |
| Preoperative NBM volume (mm²) | 0.9551 (0.8999; 1.0103), p=0.0000 |
| Age (years) | 0.2735 (-1.4613; 2.0082), p=0.7558 |
| MMSE (points) | 5.1665 (-1.7226; 12.0555), p=0.1404 |
| Female sex | 1.8579 (-15.9173; 19.6332), p=0.8366 |
| Postoperative BuChE change (U/L) | 0.0145 (-0.0052; 0.0341), p=0.1479 |
| POCD** | 6.9999 (-18.1286; 32.1284), p=0.5828 |
| Interaction between postoperative BuChE change (U/L) and POCD | 0.0016 (-0.0379; 0.0412), p=0.9357 |
| N*** | 153 |
| $R^2$ | 0.9146 |

* Intercept for: Mean preoperative NBM volume, age of 75 years, MMSE of 29p, male sex, no POCD

** POCD, postoperative cognitive dysfunction after three months

*** two patients have missing values for POCD assessment
